# Cost Benefit Analysis of Limited Reopening Relative to a Herd Immunity Strategy or Shelter in Place for SARS-CoV-2 in the United States

**DOI:** 10.1101/2020.06.26.20141044

**Authors:** Robert B. Schonberger, Yair J. Listokin, Ian Ayres, Reza Yaesoubi, Zachary R. Shelley

**Affiliations:** Associate Professor; Department of Anesthesiology; Yale School of Medicine; New Haven, CT; Shibley Family Fund Professor of Law; Yale Law School; New Haven, CT; William K. Townsend Professor of Law; Yale Law School; New Haven, CT; Assistant Professor: Yale School of Public Health; New Haven, CT; Research Fellow; Yale Law School; New Haven, CT

**Author notes:** **Corresponding Author:** Robert B. Schonberger, Department of Anesthesiology, Yale School of Medicine, 333 Cedar Street; TMP-3, New Haven, CT 06520.

## Abstract

**Background:** Fierce debate about the health and financial tradeoffs presented by different COVID-19 pandemic mitigation strategies highlights the need for rigorous quantitative evaluation of policy options.

**Objective:** To quantify the economic value of the costs and benefits of a policy of continued limited reopening with social distancing relative to alternative COVID-19 response strategies in the United States.

**Design:** We estimate the number and value of quality-adjusted life-years (QALY) gained from mortality averted, with a value of $125,000 per QALY, and compare these benefits to the associated costs in terms of plausible effects on US GDP under a policy of continued limited reopening with social distancing relative to a policy of full reopening toward herd immunity. Using the same QALY value assumptions, we further evaluate cost-effectiveness of a return to Shelter-in-Place relative to a policy of limited reopening.

**Setting:** United States

**Measurements:** QALY and cost as percent of GDP of limited reopening with continued social distancing relative to a strategy of full reopening aimed at achieving herd immunity; a limited reopening “budget” measured in the number of months before this strategy fails to demonstrate cost-effectiveness relative to a full reopening; a shelter-in-place “threshold” measured in the number of lives saved at which a month of sheltering in place demonstrates cost effectiveness relative to the limited reopening strategy.

**Results:** QALY benefits from mortality averted by continued social distancing and limited reopening relative to a policy of full reopening exceed projected GDP costs if an effective vaccine or therapeutic can be developed within 11.1 months from late May 2020. White House vaccine projections fall within this date, supporting a partial reopening strategy. One month of shelter-in-place restrictions provides QALY benefits from averted mortality that exceed the associated GDP costs relative to limited reopening if the restrictions prevent at least 154,586 additional COVID-19 deaths over the course of the pandemic. Current models of disease progression suggest that limited reopening will not cause this many additional deaths, again supporting a limited reopening strategy.

**Limitation:** Limited horizon of COVID-19 mortality projections; infection fatality ratio stable across strategies, ignoring both the potential for ICU overload to increase mortality and the deployment of partially effective therapeutics to decrease mortality; effect on GDP modeled as constant within a given phase of the pandemic; accounts for age and sex distribution of QALYs, but not effect of comorbidities; only considers impact from QALY lost due to mortality and from changes in GDP, excluding numerous other considerations, such as non-fatal COVID-19 morbidity, reduced quality of life caused by prolonged social distancing, or educational regression associated with prolonged school closures and restrictions.

**Conclusions:** A limited reopening to achieve partial mitigation of COVID-19 is cost effective relative to a full reopening if an effective therapeutic or vaccine can be deployed within 11.1 months of late May 2020. One additional month of shelter-in-place restrictions should only be imposed if it saves at least 154,586 lives per month before the development of an effective therapeutic or vaccine relative to limited reopening.

**Funding:** This work was supported in part by grant K01AI119603 from the National Institute of Allergy and Infectious Diseases (NIAID). This work does not necessarily represent the opinions of the NIAID, the NIH, or the United States Government.

## Background

As the national response to the COVID-19 pandemic evolves, the difficult health and financial tradeoffs posed by prolonged public health restrictions have grown increasingly stark. In this context, we quantify the value of Quality Adjusted Life Years (QALY) saved (or lost) by a limited reopening with continued social distancing as compared to both a pandemic scenario that ends with “herd immunity” and a return to shelter-in-place. We compare these benefits to the associated costs in terms of plausible effects on US GDP.

## Objective

We provide estimates of the number of months, beyond May 26, 2020, that the strategy of limited economic and social reopening present on that date demonstrates cost-effectiveness relative to full reopening. We also provide a threshold number of lives saved at which one month of shelter-in-place limitations would demonstrate cost-effectiveness relative to the limited reopening strategy.

## Methods and Findings

We use cost benefit analysis (1) to compare the economic cost of a continuation of limited reopening with social distancing at levels that were present on May 26, 2020, with the economic value of mortality averted by this policy as compared to a pandemic ending in late January 2021 with herd immunity. We also provide a cost benefit analysis of shelter-in-place restrictions relative to a limited economic and social reopening. Our analysis adheres to the Consolidated Health Economic Evaluation Reporting Standards (CHEERS)(2) guidelines (see online appendix B). The two strategies to which limited reopening is compared reflect both a more and a less restrictive policy alternative to the level of limited reopening that was in place on May 26, 2020.

We estimate that the mortality benefits of limited reopening versus a full reopening exceed their costs so long as such policies are not in place for more than 11.1 months (slightly less than 1 year) before the discovery and implementation of an effective vaccine or therapeutic. The estimate of a limited reopening “budget” allows for consideration of policy alternatives in light of the reader’s own estimates regarding the timing of these developments. By contrast, existing cost effectiveness analyses of COVID-19 interventions use fixed end dates.(3, 4) Such analyses implicitly assume that a vaccine will be available by the given end date. If a vaccine is not available on the ad hoc end date, however, then the public health restrictions under study have simply delayed COVID-19 mortality rather than prevented it, radically changing the cost-effectiveness evaluation.

The 11.1-month “limited reopening budget” (before limited reopening fails to demonstrate cost-effectiveness) depends, of course, on many assumptions, which we list in Table 1 and describe below. Results are shown in Table 2 and Figure 1, where we plot the cost-effectiveness of limited reopening versus full reopening under varying assumptions. At this site (hyperlink to https://covid19-cost-benefit.shinyapps.io/Covid19-Cost-Benefit/), we allow the user to insert alternative assumptions and track changes in the limited reopening “budget”, enabling the user to conduct their own sensitivity analysis. We provide further elaboration of the limited reopening “budget” concept in the discussion below.

**Table 1:**
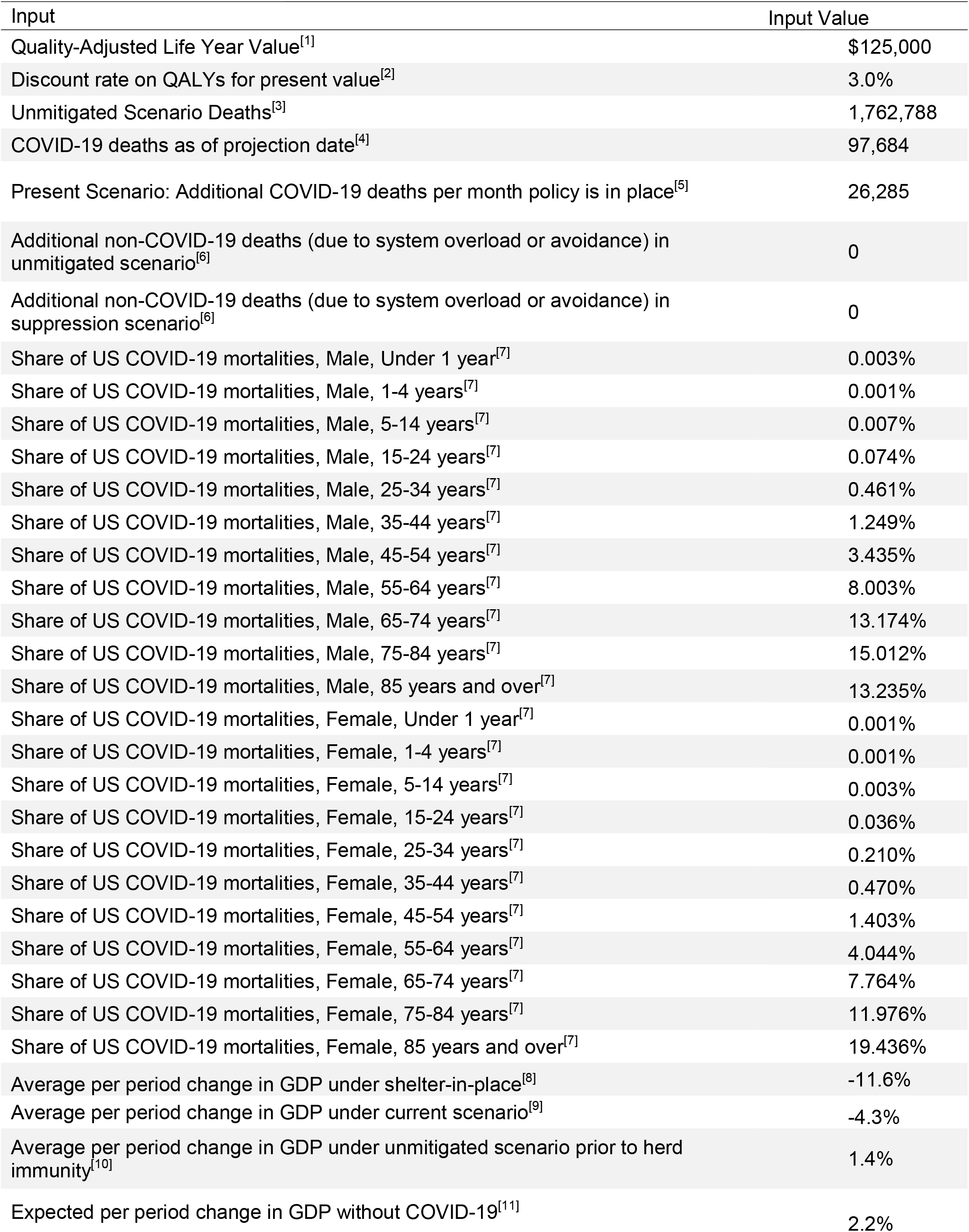
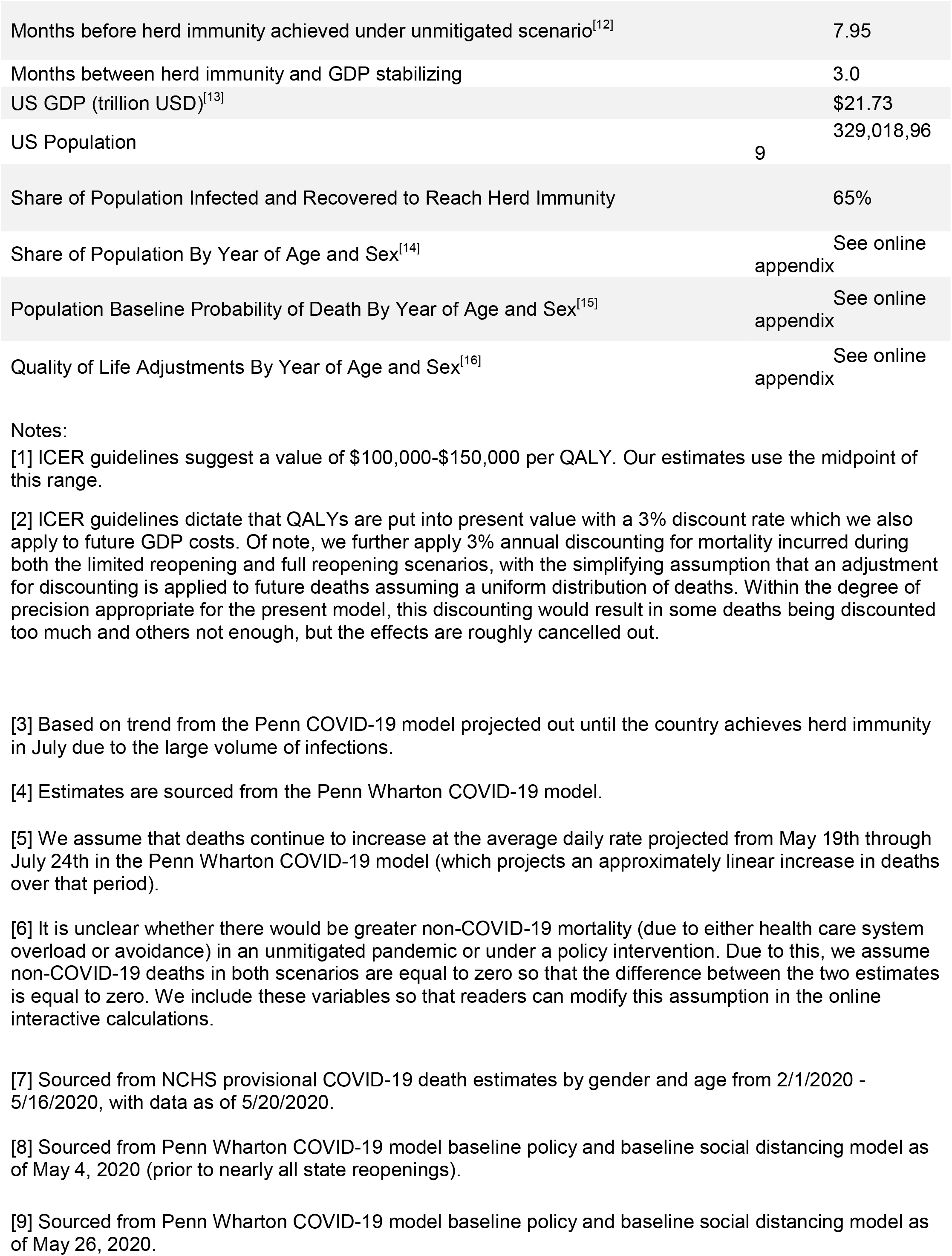
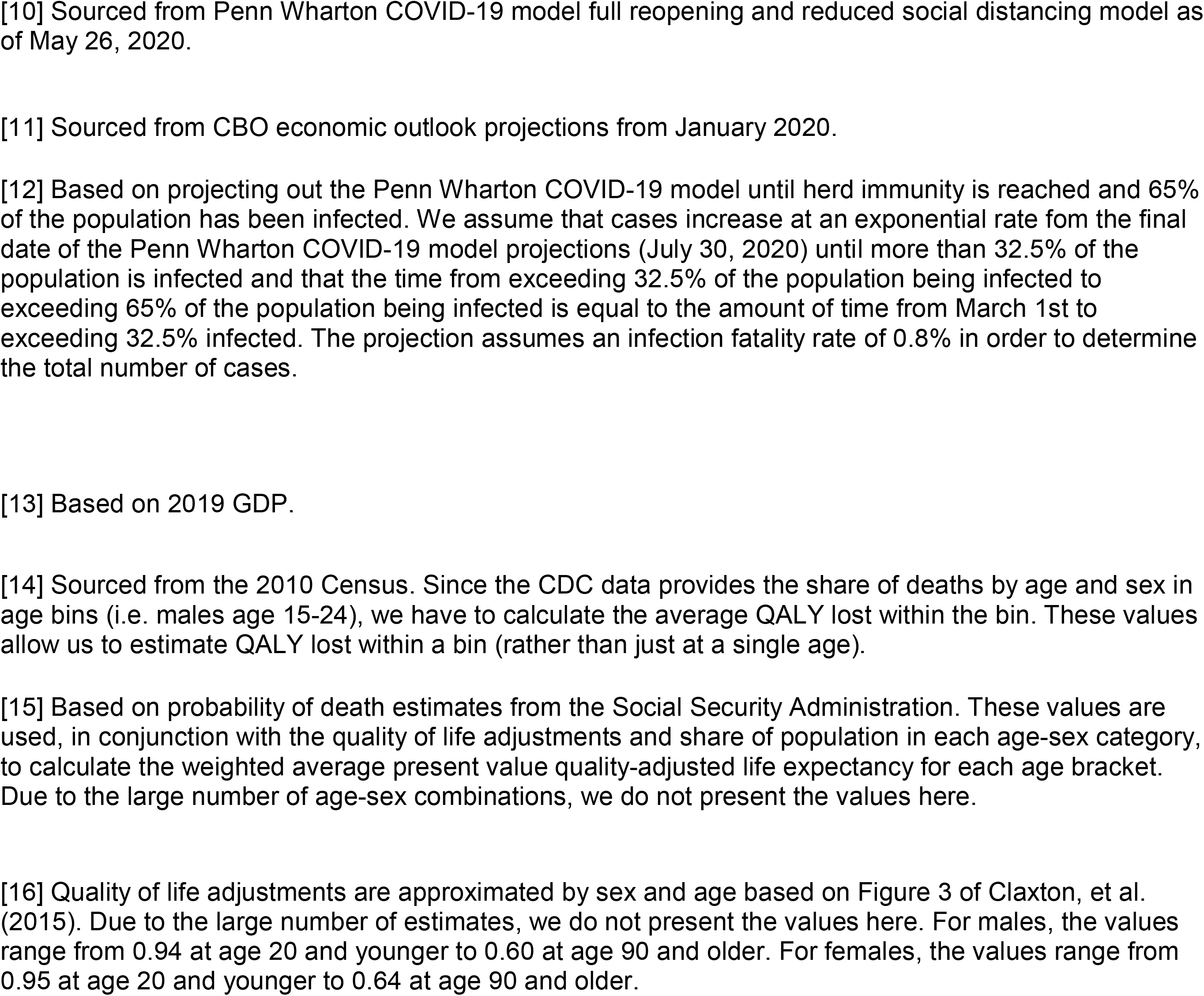
List of Assumptions

**Table 2:**
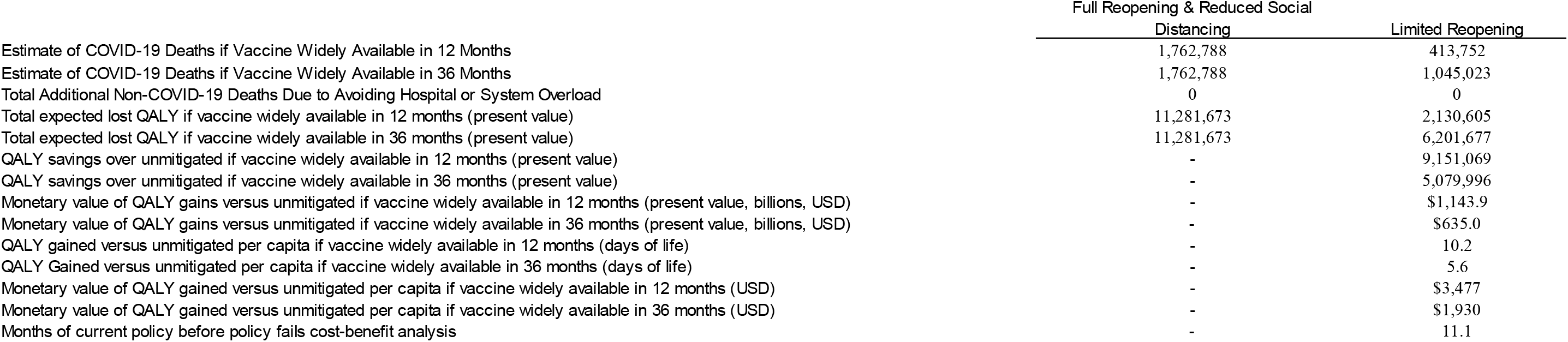
Model Results Between Full Reopening and Limited Reopening Scenarios

**Figure 1:**
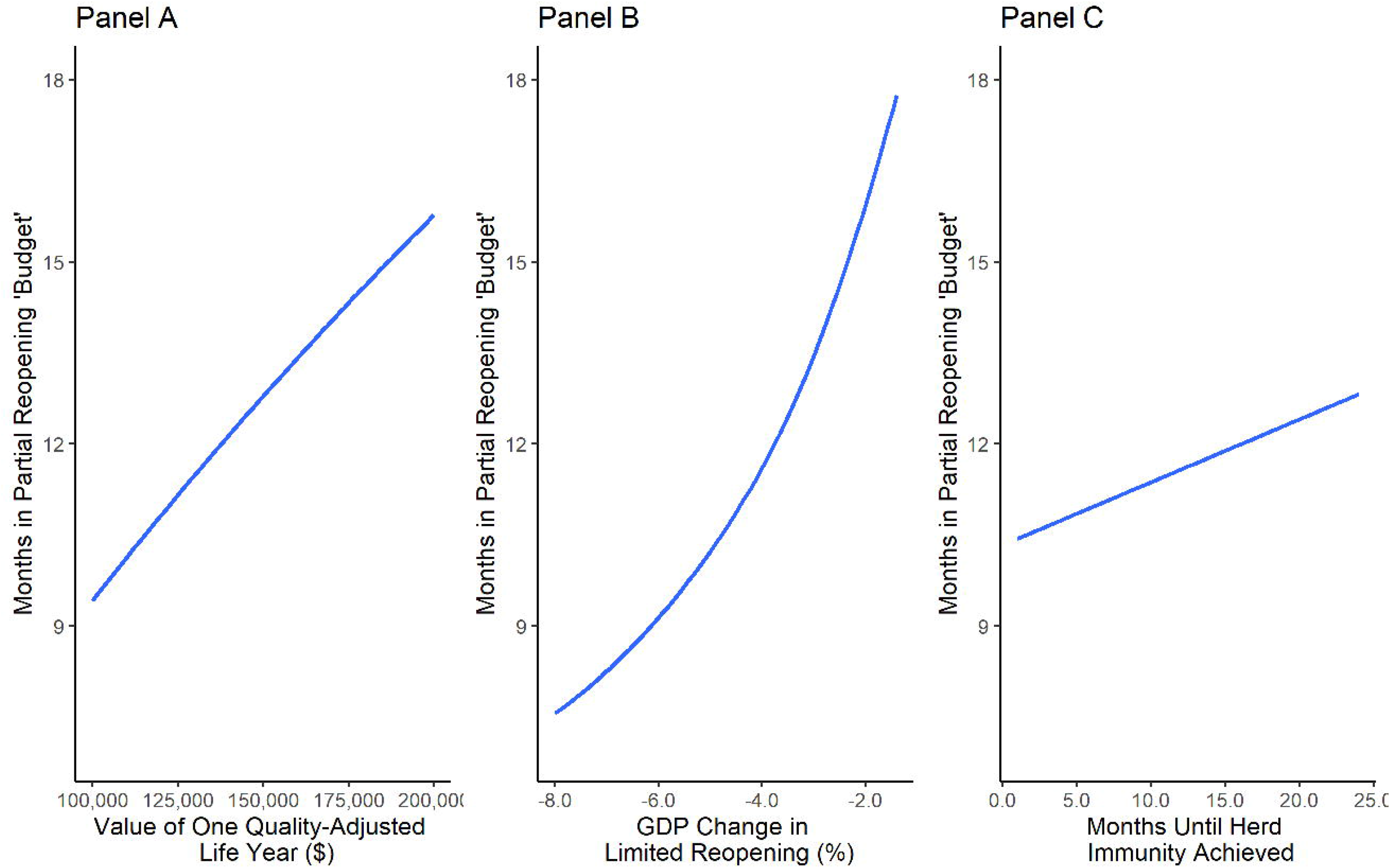
Maximum Number of Months That Limited Reopening Demonstrates Cost-Effectiveness Vs. Herd Immunity Under Changing Model Assumptions.

Using the same assumptions regarding QALY value, we find that shelter-in-place restrictions are unlikely to demonstrate cost-effectiveness relative to a limited reopening strategy. The economic harms associated with shelter-in-place exceed the value of QALY benefits unless one additional month of shelter-in-place restrictions would prevent more than 154,586 deaths, both during that month and in subsequent time periods before an effective vaccine or therapeutic is deployed. Most models, however, project that added mortality from limited opening falls well below this “shelter-in-place threshold”. Even if shelter-in-place restrictions prevent more than 154,586 deaths in a particular month, if the restrictions merely delay the deaths rather than completely preventing them through the time that a vaccine is deployed, then the additional restrictions fail cost-benefit analysis.

## Assumptions

We assume that recovery from COVID-19 confers total or partial immunity for a long-enough period to develop an effective vaccine or therapeutic before the disease spreads again. We further assume that in a herd immunity strategy, pandemic COVID-19 mortality would be 1,762,788. This number is derived by extending the Penn Wharton epidemiological model (5) “Full Reopening” policy scenario and “Reduced Social Distancing” behavior scenario under an exponential model until herd immunity is achieved (when the total number of people who have been infected and recovered) exceeds 65% of the population). In this model, herd immunity is achieved on January 23, 2021. The herd immunity mortality number is broadly consistent with pandemic mortality estimates from prior models.(6) This estimate of mortality corresponds to a 0.8% infection fatality rate for the U.S. population, falling within the range of published infection fatality rate estimates.(7, 8) If pandemic mortality can be reduced through alternative methods of achieving herd immunity, such as isolating the elderly and others at high risk from COVID-19,(9) then the partial reopening budget goes down. If our pandemic mortality is an underestimate (e.g. due to ICU overload or due to “overshooting” of herd immunity infection rates),(10) then the partial reopening budget rises. We include a figure in the online appendix that plots the relationship between pandemic mortality and the partial reopening budget (see appendix A).

## Benefits of a partial reopening regime

COVID-19 deaths associated with continued social distancing policies are obtained from the Penn Wharton epidemiological model(5) assumptions as of May 26, 2020, when the model’s “baseline” projections accounted for the maintenance of the limited reopening strategy that was in place on that date (see Table 2). We utilize the mortality projections from this model through July 2020 and assume that under the partial reopening occurring as of end of May 2020, the U.S. would limit additional COVID-19 deaths to 26,285 per month (315,420 per year), which is equal to the average monthly deaths in the Penn baseline model from May 26, 2020 through July 30, 2020. In our extension of the Penn Wharton model, mortality does not flatten further but merely continues at this roughly linear pace as the search for a vaccine or therapeutic continues. Maintaining a limited reopening policy that is restricted to the May 26, 2020 level over the next 12 months therefore saves approximately 1.35 million lives relative to a relatively unrestrained pandemic ending in herd immunity, or on the order of approximately 9.1 million quality adjusted life-years saved relative to full reopening. If a vaccine or therapeutic is not deployed until 36 months from late May 2020, maintaining a limited reopening would save 718,000 lives (5.0 million QALY). These QALY estimate reflect the age- and sex-adjusted lost QALY expectancy of COVID-19 victims. If true mortality from an alternative herd immunity response is substantially higher or lower than this modeled estimate, then the limited reopening budget rises or falls accordingly.

We estimate the economic value of the quality adjusted life years (QALYs) saved by limited reopening relative to a full reopening using the Institute for Clinical and Economic Review (ICER) value assessments framework(11) with a QALY value of $125,000 and a discount rate of 3% (standard assumptions), in concert with Social Security Administration actuarial life tables,(12) age and sex distributions of the population based on the 2010 Census,(13) plausible age and sex distributions of COVID-19 mortality based on Centers for Disease Control and Prevention data,(14) and estimates of quality of life scores from previous literature.(15) Alternative values for a QALY can be chosen at the reader’s discretion using our online calculator, and the effects of changes in QALY valuation are illustrated in Figure 1 Panel A

Because COVID-19 mortality is skewed toward males and the elderly, the QALY framework reduces the economic benefit of lives saved relative to using a fixed statistical value per life and therefore shortens the limited reopening budget relative to analyses that refer only to mortality totals.(16) We do not adjust for reduced QALY from COVID-19 morbidity among those who recover from the disease (which would lengthen the limited reopening budget) or from reduced QALY expectancy associated with pre-existing COVID-19 risk factors and comorbidities such as obesity, hypertension, and diabetes (which would shorten the limited reopening budget).(17)

Relative to shelter-in-place restrictions, the benefits of limited reopening are economic. Following the Penn-Wharton model, we estimate that GDP per period is 7.3% higher year-on-year under limited reopening than under shelter-in-place.

## Costs of continuing a partial reopening regime

We rely on the Penn Wharton integrated economic and epidemiological model to approximate the economic costs of a limited reopening in comparison to a full reopening, herd immunity strategy.(5) The Penn Wharton model was chosen as the most prominent model of which we are aware that provides both mortality estimates and year over year effects of alternative strategies on GDP. In this model, maintaining a limited reopening (baseline policy and baseline behavior scenarios in the Penn Wharton model as of May 26, 2020) reduces GDP by 4.3% measured year-on-year (i.e. May 2020 GDP will be 4.3% lower than May 2019 GDP). Lesser restrictions (“full reopening” policy and “reduced social distancing” behavior scenarios) in the Penn Wharton model are associated with an increase in GDP of 1.4% year on year, an absolute decrease of 0.8% from the Congressional Budget Office’s pre-COVID projections of 2.2% growth.(18) Of note, the Penn Wharton model attributes less economic importance to individual behavioral changes that are independent of official pronouncements than prior modeling of the economic effects of pandemics.(18-20) All GDP calculations are discounted at 3% annually in our calculations in parallel with QALY discounting. Alternative assumptions regarding the GDP cost of limited reopening on GDP are illustrated in Figure1 Panel B.

Following the hypothetical achievement of herd immunity at 65% of the population recovered from infection as of January 23, 2021, we assume that the Penn Wharton projection of GDP effects slowly improves, returning to 2019 levels of 2.2% growth three months after the pandemic ends as economic uncertainty resolves and activity restrictions and voluntary social distancing slowly end. A longer or shorter period of time prior to achievement of herd immunity and its effects on our partial reopening budget is illustrated in Figure 1, Panel C.

We assume that non-economic costs associated with partial reopening are $0. For example, our model includes no cost if educators are paid and students attend school remotely, even if student learning suffers. Including important non-economic costs such as the value of lost learning,(21) would reduce the length of the limited reopening budget. Users of the online tool can simulate these effects by increasing the GDP cost of limited reopening above 4.3%. Figure 1 Panel C plots the limited reopening budget across a variety of GDP costs.

Relative to a shelter-in-place strategy, limited reopening is associated with higher mortality. The cost of limited reopening relative to shelter-in-place is the value assigned to the QALYs saved by shelter-in-place. Our shelter-in-place threshold provides the number of lives that must be saved by a month of shelter in place *relative* to limited reopening to justify returning to shelter-in-place. If shelter-in-place restrictions do not avoid fatalities but rather delay them to later periods, then these delayed fatalities are not included in the shelter-in-place threshold. Instead, an additional month of shelter-in-place needs to save at least the threshold number of lives until the deployment of an effective vaccine or therapeutic.

Our simulation assumes there is no difference in non-COVID-19 deaths between the pandemic scenario and the limited reopening scenario, so long as all deaths from COVID-19 are properly counted. We recognize that the pandemic is causing non-COVID-19 mortality to rise substantially due to both a lack of hospital treatment capacity and avoidance and delay of medical care. Because the pandemic spreads relatively slowly even under “full reopening with reduced social distancing” in the Penn Wharton model, the danger of overwhelming hospitals appears lower than earlier fears. With respect to avoidance and delay of medical care, the pandemic ends within 8 months. After this point, we assume that patients access medical care at normal rates. If the wait for a widely available vaccine takes longer than 8 months, limited reopening may therefore be associated with more avoidance and delay of care than a pandemic. Rather than making an uninformed guess about the size of these effects, we allow readers to enter their own estimates in the online tool. If non-COVID-19 mortality is in fact higher from ICU overload or other effects in the pandemic scenario, then the limited reopening budget reported above should be longer.

## Discussion

COVID-19 presents horrific policy choices. The restrictions necessary to avoid unprecedented disease mortality impose unprecedented economic calamity. Our cost benefit analysis attempts to compare these factors using a widely used framework. Given current expectations about future vaccine availability, we conclude that maintaining limited reopening with social distancing to the levels present on May 26, 2020 demonstrates cost effectiveness relative to both a policy of shelter in place and a policy of full reopening toward herd immunity. As additional information on a vaccine timeline, the non-economic costs of partial reopening, and the potential increase in mortality due to ICU overload in a relatively unrestrained pandemic become available, this conclusion may change. Our online tool allows policymakers to reassess the net effect of these tradeoffs as new information appears.

We estimate that shelter in place restrictions need to prevent at least 154,586 COVID-19 deaths for each month they are in effect to demonstrate cost effectiveness relative to limited reopening, a threshold which is unlikely to be met. While readers may experience this calculus favoring a limited reopening over a return to shelter in place as unduly grisly, we would suggest that the healthcare community, focused on patient care, may (understandably) skew toward a perspective that emphasizes the costs of lives lost to disease over the costs of unemployment rivaling the Great Depression and dislocated family and educational lives. However, the disparities in how the socio-economic burden is imposed upon the most disadvantaged relative to the well-off may be even more inequitable than the disparities in health outcomes with which the healthcare community is so familiar.

11.1 months (or a revised estimate derived from different assumptions) is our “limited reopening budget” for mitigating COVID-19. The limited reopening budget means that if we think, as of the end of May 2020, that we can implement an effective COVID-19 therapy or vaccine within 11.1 months, as the White House has publicly proclaimed, then we should pursue a coordinated national strategy to maintain a limited reopening at the levels in place as of May 26, 2020, and accept the grievous economic consequences. Alternatively, if we find ourselves unsure regarding treatment and vaccine timetables, then we can justify temporarily maintaining limited reopening to push off both the large short-term mortality costs of full reopening and the economic costs of shelter-in-place restrictions. If we expect the waiting period to resolve the uncertainty around vaccine time-tables, this delay would prevent unnecessary death and economic harm.(22)

If instead we conclude that these scenarios are wishful thinking and we face an expected wait time longer than 11.1 months, then we should develop the most effective “herd immunity” strategy we can (such as concentrating infection among those below age 50) and implement it to minimize economic devastation and loss of life. Picking any one of these devastating health and economic choices is, and should be, repellent, but such is the dilemma presented by COVID-19.

## Data Availability

Files to reproduce models will be made public upon peer review.

https://covid19-cost-benefit.shinyapps.io/Covid19-Cost-Benefit/

## References

1. Boardman A, Greenberg D, Vining A, Weimer D. Cost-Benefit Analysis: Concepts and Practice; 4th edition; New York: Cambridge University Press; 2018.

2. Husereau D, Drummond M, Petrou S, al. E. Consolidated health economic evaluation reporting standards (CHEERS) - explanation and elaboration: a report of the ISPOR Health Economic Evaluations Publication Guidelines Good Reporting Practices Task Force. Value Health. 2013;16(2):231–50.

3. Thunström L, Newbold SC, Finnoff D, Ashworth M, Shogren JF. The Benefits and Costs of Using Social Distancing to Flatten the Curve for COVID-19. Journal of Benefit-Cost Analysis. 2020:1–17.

4. Greenstone M, Nigam V. Dose Social Distancing Matter?; University of Chicago, Becker Friedman Institute for Economics Working Paper No. 2020-26 (url: https://papers.ssrn.com/sol3/papers.cfm?abstract_id=3561244; as accessed June 1, 2020..

5. Wharton School of Business; Coronavirus Policy Response Simulator: Health and Economic Effects of State Reopenings; url: https://budgetmodel.wharton.upenn.edu/issues/2020/5/1/coronavirus-reopening-simulator as accessed May 4, 2020.

6. Ferguson NM, Laydon D, Nedjati-Gilani G, Natsuko I, Ainslie K, Baguelin M, et al. Impact of non-pharmaceutical interventions (NPIs) to reduce COVID-19 mortality and healthcare demand; url: https://www.imperial.ac.uk/media/imperial-college/medicine/sph/ide/gida-fellowships/Imperial-College-COVID19-NPI-modelling-16-03-2020.pdf as accessed March 23, 2020.

7. Russell TW, Hellewell J, Jarvis CI, van Zandvoort K, Abbott S, Ratnayake R, et al. Estimating the infection and case fatality ratio for coronavirus disease (COVID-19) using age-adjusted data from the outbreak on the Diamond Princess cruise ship, February 2020. Euro Surveill. 2020;25(12).

8. Verity R, Okell LC, Dorigatti I, Winskill P, Whittaker C, Imai N, et al. Estimates of the severity of coronavirus disease 2019: a model-based analysis. The Lancet Infectious Diseases. 2020.

9. Acemoglu D, Chernozhukov V, Werning I, Whinston MD. A Multi-Risk SIR Model with Optimally Targeted Lockdown; National Bureau of Economic Research; No. 27102; url: https://www.nber.org/papers/w27102.pdf as accessed May 5, 2020.

10. Bergstrom CT, Dean N. What the Proponents of ‘Natural’ Herd Immunity Don’t Say; New York Times; May 1, 2020; url: https://www.nytimes.com/2020/05/01/opinion/sunday/coronavirus-herd-immunity.html as accessed May 5, 2020.

11. Weiss RD, Potter JS, Griffin ML, Provost SE, Fitzmaurice GM, McDermott KA, et al. Long-term outcomes from the National Drug Abuse Treatment Clinical Trials Network Prescription Opioid Addiction Treatment Study. Drug & Alcohol Dependence. 2015;150:112–9.

12. Social Security Administration: Actuarial Life Table 2015; url: https://www.ssa.gov/oact/STATS/table4c6_2015.html as accessed May 5, 2020.

13. Howden JM, Meyer JA. Census Briefs: Age and Sex Composition: 2010; United States Census Bureau; US Department of Commerce (2011 May); url: https://www.census.gov/prod/cen2010/briefs/c2010br-03.pdf as accessed May 5, 2020.

14. Coronavirus 2019 (COVID-19) Surveillance Data; National Vital Statistics System, National Center for Health Statistics. url: https://www.cdc.gov/nchs/nvss/vsrr/covid19/index.htm as accessed May 2, 2020.

15. Claxton K, Martin S, Soares M, al. E. Methods for the estimation of the National Institute for Health and Care Excellence cost-effectiveness threshold. Southampton (UK): NIHR Journals Library; 2015 Feb. (Health Technology Assessment, No. 19.14.) Appendix 3, Translating mortality effects into life-years and quality-adjusted life-years; url: https://www.ncbi.nlm.nih.gov/books/NBK274321/ as accessed May 5, 2020.

16. Hammitt JK. QALYs Versus WTP. Risk Analysis. 2002;22(5):985–1001.

17. Richardson S, Hirsch JS, Narasimhan M, Crawford JM, McGinn T, Davidson KW, et al. Presenting Characteristics, Comorbidities, and Outcomes Among 5700 Patients Hospitalized With COVID-19 in the New York City Area. JAMA. 2020.

18. Keogh-Brown MR. The possible macroeconomic impact on the UK of an influenza pandemic. Health economics. 2010;19(11):1345–60.

19. Dandekar R, G. B. Quantifying the effect of quarantine control in Covid-19 infectious spread using machine learning; medRxiv; url: https://www.medrxiv.org/content/10.1101/2020.04.03.20052084v1 as accessed April 21, 2020.

20. Friedson AI, McNichols D, Sabia JJ, Dave D. Did California’s Shelter-in-Place Order Work? Early Coronavirus-Related Public Health Effects; National Bureau of Economic Research Working Paper Series: No. 26992; url: http://www.nber.org/papers/w26992 as accessed April 22, 2020.

21. Soland J, Kuhfeld M, Tarasawa B, Johnson A, Ruzek E, Liu J. The Impact of COVID-19 on student achievement and what it may mean for educators; Brown Center Chalkboard; Brookings Insitute; May 27, 2020. url:https://www.brookings.edu/blog/brown-center-chalkboard/2020/05/27/the-impact-of-covid-19-on-student-achievement-and-what-it-may-mean-for-educators/; as accessed June 1, 2020.

22. Ayres I, Listokin YJ, Schonberger RB. The Option Preserving Value of Social Distancing; Incidental Economist, url: https://theincidentaleconomist.com/wordpress/the-option-preserving-value-of-social-distancing/ as accessed April 24, 2020.

